# NI BabyHearts 2 study protocol: Examining preconceptional and antenatal factors in relation to developmental and educational outcomes of children with and without congenital heart disease

**DOI:** 10.64898/2026.09.02.26362009

**Authors:** Lesley-Anne Henry, Helen Dolk, Joanne Given, Maria Loane, Victoria Simms, Paul Slater, Frank Casey

## Abstract

**Background:** Congenital heart disease is the most common congenital anomaly and a major public health concern. No single cause has been identified, and its aetiology is most likely to be multifactorial, determined by a combination of genetic and environmental factors. Congenital heart disease can have a significant and lasting impact on children’s health and development with well documented neurodevelopmental difficulties experienced by some children with CHD. However, evidence linking early environmental factors to developmental outcomes remains limited, despite growing recognition of their importance. The Northern Ireland BabyHearts 2 Study provides an opportunity to investigate these relationships in a longitudinal cohort with extensive prospectively collected preconceptional and antenatal data.

**Aim:** To investigate the associations between preconceptional and antenatal environmental factors and the birth outcomes, physical health, neurodevelopment and academic achievement of children with and without congenital heart disease.

**Methods:** This is a longitudinal matched cohort follow-up study of children with and without congenital heart disease recruited to the NI BabyHearts 1 case-control study (2014–2017), with controls matched 1:1 by age and sex. Data sources include the existing NI BabyHearts 1 dataset of prospectively collected preconceptional and antenatal environmental exposures, together with linked routine maternity and prescribing data. Direct follow-up data collected at 8-11 years of age included parental questionnaires assessing child executive function and behavioural development, standardised child-completed assessments of neurodevelopment, language and educational attainment, and clinical information.

**Impact:** This study will provide evidence on the long-term health and developmental outcomes in children born with and without congenital heart disease and improve understanding of how early environmental factors influence these outcomes. Findings will inform clinical care and service provision for children with congenital heart disease, strengthen preconception and antenatal guidance, and contribute to public health policy aimed at improving long-term outcomes for children living with congenital heart disease.

## Introduction

Congenital Heart Disease (CHD) is the most common birth defect in children accounting for almost one third of congenital anomalies diagnosed prenatally or in infancy [1]. The average global prevalence is 8 per 1,000 live births, equating to approximately 1.35 million babies born worldwide each year with some form of CHD [2,3]. CHD is considered a major public health concern and global burden [4] and is a leading cause of infant morbidity and mortality. The treatment of CHD is one of the success stories of modern medicine with surgical advances and improved postoperative care tremendously increasing survival rates. More than 90% of patients are now expected to reach adulthood [5].

No single cause of CHD has been identified, and, in most cases, it is likely to be multifactorial. Similarly, developmental outcomes among children with CHD are influenced by a complex combination of genetic, antenatal, postnatal and clinical factors (*Fig 1*). While genetic contributions are increasingly understood, the role of maternal environmental risk and protective factors requires further investigation [6]. Cardiac morphogenesis occurs during early pregnancy (8–10 gestational weeks), making this a critical window of vulnerability [7,8]. Similarly, brain development begins at conception, with rapid cortical maturation during the third trimester [9–11]. Abnormal fetal circulation associated with CHD can also affect brain development and maturation. Environmental exposures during these critical periods may therefore influence later neurodevelopmental outcomes. Postnatal refinement of neuronal pathways through synaptic plasticity further highlights the continuing influence of lived experience and environmental factors on human development [12].

**Fig 1:**
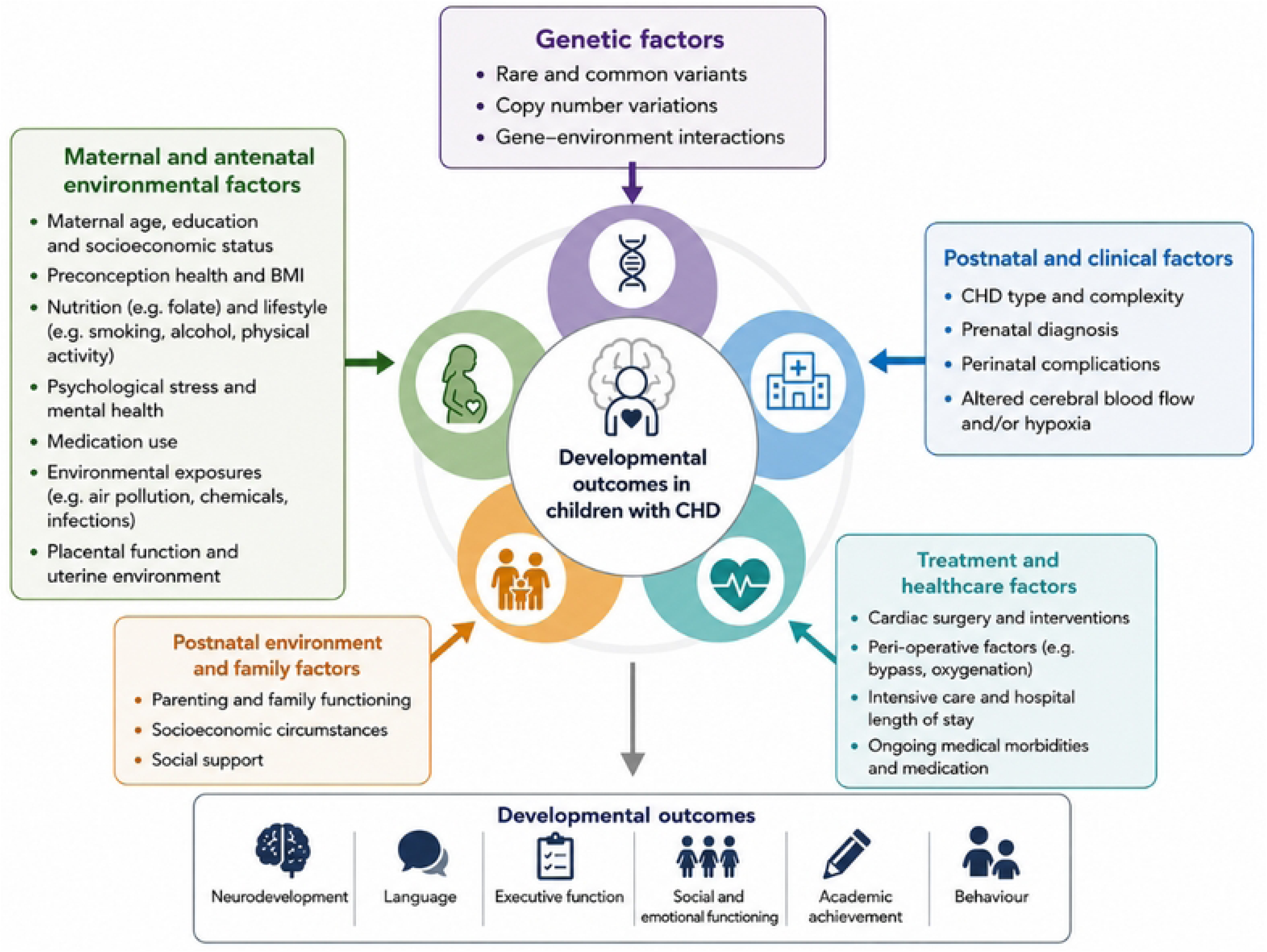
Factors influencing developmental outcomes in children with congenital heart disease.

### Early Northern Ireland BabyHearts Investigations

The Northern Ireland (NI) BabyHearts 1 Study (BH1) was conducted between 2014–2017 and is described in detail elsewhere [6,13]. Briefly, BH1 investigated possible common environmental risk and protective factors for CHD in 242 cases and 966 controls using parent completed questionnaires, self-completed on an iPad during antenatal or paediatric cardiology clinic visits, together with administrative data linkage. Several maternal environmental factors were found to be independently associated with CHD risk, including, sociodemographic factors (maternal education), maternal medical conditions (pregestational diabetes, clotting disorders and their treatment, vaginal infections), maternal mental health particularly stress, and various dietary factors [13].

The extensive data collected during BH1 created a rich database of maternal exposures present during pregnancy, which can now be examined in relation to later developmental outcomes collected during the current study. A full BH1 variable list is available (S1 *Appendix*), in summary the NI BabyHearts dataset contains sociodemographic characteristics, lifestyle information (e.g. smoking, alcohol, diet, exercise), maternal and paternal mental health and stressors, maternal and paternal medical history, and maternal prescriptions from preconception to 12 gestational weeks (GW). Clinical data regarding children’s CHD diagnosis is also included.

### CHD and Later Childhood Development

The impact of CHD on children’s health outcomes has been subject to much investigation, with CHD type, severity and complexity having been established as contributors to these children’s outcomes. In addition, perioperative factors, including cardiac surgery, the timing of surgical intervention and postoperative complications, have also been implicated in neurodevelopmental outcomes [14–17]. Aside from impacting children’s physical [18–21] and mental health [22,23], CHD has been associated with adverse neurodevelopmental and cognitive outcomes in some children [5,24–28] as well as difficulties in psychological development [29–32]. Although difficulties appear to be more prevalent and severe in those with complex heart conditions, those with less severe or uncorrected CHD remain at increased risk of comorbidities and are comparatively understudied [28,33]. Environmental and family factors, including family functioning and parenting approaches, may also influence children’s outcomes and, in some contexts, predict outcomes more strongly than biological factors alone [34,35].

### Antenatal factors and Neurodevelopment

The neurodevelopmental difficulties experienced by some children with CHD are well documented. Despite this, the contribution of maternal, pregnancy-related and socioeconomic factors to longer-term neurodevelopmental outcomes in this population remains comparatively underexplored. These factors are already recognised as important in child development generally, so understanding their role in children with CHD is an important next step. A recent scoping review [36] found that maternal education and SES were among the most consistently examined factors associated with developmental outcomes in children with CHD, while maternal health and pregnancy-related factors were less frequently investigated. The review also highlighted the limited number of prospective longitudinal studies examining maternal and environmental influences on longer-term neurodevelopmental outcomes in children with CHD.

### The Current Study – NI BabyHearts 2

BH1 collected detailed information on a wide range of maternal, pregnancy-related, nutritional and socioeconomic factors, including many of the exposures identified in the literature as potentially relevant to neurodevelopmental outcomes in children with CHD. Within the BH1 cohort, several maternal environmental factors were associated with CHD risk, including maternal stress, low maternal education and dietary factors [13]; however, the potential longer-term influence of these and other early-life environmental factors on child development has not yet been examined. NI BabyHearts 2 (BH2) provides a unique opportunity to extend this work by linking prospectively collected maternal and antenatal data with neurodevelopmental outcomes in later childhood. As some of these antenatal factors are also associated with poorer neurodevelopmental outcomes in children without the condition, the BH1 control group also provides an opportunity to analyse whether children with CHD are particularly vulnerable to certain environmental factors, or whether their vulnerability mirrors that of children without CHD.

### Aim and Objectives

*Aim:* To investigate the associations between preconceptional and antenatal environmental factors and birth outcomes, physical health, neurodevelopment and academic achievement in NI BabyHearts children (8-11y), born with and without CHD.

*Objectives:*

1. Assess neurodevelopmental, psychological and educational outcomes in children with CHD compared with normative reference data.
2. Assess the associations between preconceptional and antenatal risk factors and perinatal birth outcomes in children with and without CHD from the NI BabyHearts cohort.
3. Examine the associations between preconceptional and antenatal environmental factors and neurodevelopmental, psychological and educational outcomes in NI BabyHearts children with CHD.
4. Examine associations between these factors and physical development in NI BabyHearts children.
5. Determine whether maternal factors previously identified as independent risk factors for CHD in BH1 also predict adverse developmental outcomes in children with and without CHD.
6. Compare parent-reported measures of executive function and behaviour between children with and without CHD.

## Materials and Methods

### Study Design and Participants

NI BabyHearts 2 (BH2) is a longitudinal matched cohort follow-up of participants recruited to the BH1 case-control study, designed to examine associations between early-life exposures and developmental outcomes in children with and without CHD. While the original BH1 study collected detailed preconceptional and antenatal information up to 12GW, a critical window for cardiac development, BH2 extends the exposure window from 12GW to 6 weeks post birth, a period encompassing continued brain development and important changes in neonatal haemodynamics. This extended exposure allows for a more comprehensive assessment of maternal and infant factors. The study protocol and associated study documents were preregistered and made publicly available on the Open Science Framework (OSF) on 26 November 2024 (https://doi.org/10.17605/OSF.IO/M9AS7).

### Original NI BabyHearts Study (2014-2017)

The NI BabyHearts Study 1 was a population-based case-control study conducted in Northern Ireland (NI) [6,13]. Cases were babies with CHD diagnosed prenatally or before six months of age through the single paediatric cardiology centre serving Northern Ireland.

Pregnancies where stillbirth occurred with prenatally diagnosed CHD were included. Exclusion criteria included babies born with specific confirmed abnormalities (i.e., Trisomy 21) or genetic syndromes often associated with CHD. Cases of patent ductus arteriosus associated with preterm birth, patent foramen ovale or small atrial septal defects (ASD <4mm on 2-dimensional echocardiography) were excluded.

Controls were recruited using a “one month per health unit” approach designed to achieve a representative population sample. Each of the 17 health units providing fetal anomaly scans in NI was allocated a designated recruitment month, during which all eligible women attending their routine anomaly scan at 18–22 weeks’ gestation were invited to participate. A total of 242 cases and 966 controls were recruited, with 95% consenting to future recontact.

### NI BabyHearts 2 Follow-Up (2024-2027)

BH2 follows up all surviving children with CHD who meet the eligibility criteria and a matched cohort of children without CHD from the original control group. Participant groups will be matched 1:1 by age and child’s sex. Socioeconomic Status defined using the Northern Ireland Multiple Deprivation Measure [37], will be incorporated into the analyses. This design allows associations between preconceptional and antenatal environmental factors and child outcomes to be examined while accounting for sociodemographic context. Follow-up data collection for children with and without CHD included parental questionnaires and includes ongoing linkage to administrative datasets, including the Northern Ireland Maternity System (NIMATS) and Enhanced Prescribing Database (EPD). Children with CHD additionally completed standardised developmental assessments, and clinical data were obtained from their medical records.

Participant identifiers and contact details were accessed by authorised study researchers for tracing and recruitment to BH2. Prior to participant contact, clinical records were reviewed to identify any circumstances that might make recruitment contact inappropriate or affect eligibility for follow-up, including death, a syndromic or genetic diagnosis after BH1, or other relevant clinical information. The existing BH1 research dataset was first accessed for BH2 research purposes on 2 October 2024; this dataset was pseudonymised and did not contain directly identifying information. Clinical information for children with CHD was subsequently extracted from identifiable medical records between 2 February 2026 and 29 May 2026.

As a follow-up of a fixed population-based cohort, the maximum eligible sample was determined by participation in BH1. Based on an anticipated 70% response rate, approximately 170 children with CHD and an equivalent number of matched controls were expected to participate at study outset [38,39]. In the absence of published effect-size estimates for neurodevelopmental outcomes measured using the NEPSY-II in children with CHD, estimates from a comparable study of neurodevelopment using the NEPSY-II indicated that the anticipated sample would provide sufficient power to detect small-to- moderate effects [40]. Recruitment was facilitated through collaboration with paediatric cardiology services at the Royal Belfast Hospital for Sick Children (RBHSC) and the Children’s Heartbeat Trust, a Northern Ireland based charitable organisation supporting families affected by CHD.

### Study Status

At the time of manuscript submission, participant recruitment and developmental assessments had been completed. Recruitment took place from 27 November 2024 to 1 March 2025, and developmental assessments were completed between 6 December 2024 and 6 December 2025. Data collection for the study as a whole remains ongoing, with acquisition and linkage of routine maternity and prescribing data through the Honest Broker Service (HBS) still to be completed. The final linked study dataset is therefore not yet available and the primary analyses described in this protocol have not been completed. HBS data acquisition and linkage are expected to be completed by December 2026, with the planned analyses commencing thereafter and study results expected by March 2027.

### Public and Patient Involvement and Engagement

Public and Patient Involvement and Engagement (PPIE) has been embedded from the outset of BH2. Parents of children with CHD and representatives from Children’s Heartbeat Trust have contributed to the study design, development of participant materials and recruitment strategy. The PPIE Advisory Group continues to advise throughout data collection, interpretation of findings and dissemination planning. Participant engagement has also been supported through initiatives such as the NI BabyHearts Family Appreciation Day, which provided participating families with study updates, opportunities to engage with the research team and acknowledgement of their contribution to the study. Their ongoing involvement and engagement will help ensure that the research remains relevant to the needs and priorities of children with CHD and their families and maximise the potential for findings to inform clinical practice and public health policy.

### Ethics Statement

Ethical approval was obtained from Cambridge South Research Ethics Committee on 6 March 2024 (REC Reference: 24/EE/0013; Protocol Number: 23/0082; IRAS Project ID: 334800), with research governance approval obtained from Belfast Health and Social Care Trust (BHSCT) on 20 November 2024 (HSC Trust Reference Number: 23128FC-CH).

Written informed consent was obtained from parents or legal guardians prior to completion of the survey and, where applicable, their child’s participation in the developmental assessment. Written assent was obtained from children at the beginning of the assessment session. Consent procedures cover participation in the follow-up study, access to medical records and linkage to administrative health data, as applicable. All data will be managed in accordance with approved data protection, confidentiality and information governance procedures.

## Measures and Outcomes

Additional exposure and developmental outcome data were collected using parental surveys and direct developmental assessments for children with CHD, while additional routine healthcare data are being obtained through data linkage for children with and without CHD. Clinical information was also obtained from the medical records of children with CHD. The measures and outcomes obtained from each source are described below.

### Parental Proxy Measures

Parents or guardians of both children with and without CHD completed a questionnaire designed to capture information on the child’s health, development, family, and wider environment. The questionnaire contained approximately 120 items, including filter questions, and was developed with reference to the neurodevelopmental literature on children with CHD while ensuring relevance to children more broadly. The complete questionnaire is available on the Open Science Framework (OSF) [<u>OSF | 7. Parent</u> <u>Survey.docx</u>]. It combined questions developed by the study team on parental employment and household income, parental and family chronic health conditions and associated medication use, and children’s physical health, developmental conditions, educational support, school attendance and physical activity, alongside validated parental rating scales. These included the Behaviour Rating Inventory of Executive Function, Second Edition (BRIEF-2) [41], which assesses executive functioning in children aged 5–18 years; the Strengths and Difficulties Questionnaire (SDQ) [42], which measures behavioural and emotional strengths and difficulties in children aged 2–17 years; the Brief Assessment of Family Functioning Scale (BAFFS) [43]; and an adapted version of the Parenting Styles and Dimensions Questionnaire (PSDQ) [44].

The questionnaire was administered digitally to improve efficiency, reduce completion time, and maintain confidentiality, with a paper version and prepaid return envelope available on request. To minimise recall bias and reduce participant burden, survey information will be verified and supplemented with administrative data sources such as NIMATS and the EPD, and clinical medical records for children with CHD. Parent-reported height and weight were confirmed during developmental assessments for children with CHD.

### Child-Completed Developmental Assessments (Children with CHD only)

Children with CHD completed a standardised battery of developmental assessments to comprehensively evaluate neuropsychological functioning, cognition, communication and academic achievement. Instruments included subtests from the NEuroPSYchological Assessment, Second Edition (NEPSY-II) [45], to evaluate children’s executive function, attention, memory, and learning in children aged 3–16 years; The British Ability Scales, Third Edition (BAS3) [46], to measure academic achievement in children aged 3–17 years; and the British Picture Vocabulary Scale, Third Edition (BPVS-3) [47], to assess receptive vocabulary in children aged 3–16 years. Details of the subtests administered, and the developmental domains assessed are provided in *S2 Appendix*. Anthropometric measurements, including height, weight, waist circumference and head circumference, were taken by trained researchers using standardised equipment.

This combination of measures provides an in-depth characterisation of developmental outcomes among children with CHD, and will allow comparisons by CHD severity and treatment, and enable examination of associations between antenatal exposures and developmental outcomes.

### Data Linkage

Data linkage to multiple administrative healthcare datasets is being undertaken to supplement data provided by NI BabyHearts 2 participants. Data are being accessed via the HBS and analysed within the Business Services Organisation (BSO) Safe Haven using their Secure e-Research Platform (SeRP). The Northern Ireland Maternity System (NIMATS) is a routinely collected maternity database containing demographic, maternal, pregnancy and birth information. It will be used to obtain antenatal and birth outcomes, including gestational age, birthweight, Apgar score, mode of delivery, and breastfeeding at discharge. The Enhanced Prescribing Database (EPD) is a population-based database of primary care prescriptions dispensed by community pharmacies across Northern Ireland. It will be used to extend maternal prescription data from 12GW through to birth and the first six weeks post-birth. Medications of interest will include those relating to cardiovascular, respiratory, central nervous system, endocrine and nutritional conditions, consistent with the medication categories examined in BH1 (S1 *Appendix*).

### Clinical Information (Children with CHD)

Medical records of children with CHD were reviewed using a structured proforma developed by an experienced clinician to extract information on CHD diagnosis and treatment, surgical procedures, comorbidities and clinical follow-up.

### Statistical Methods

Data will be screened for accuracy, completeness, missingness and distributional properties before the primary analyses are undertaken. Descriptive statistics will characterise children with and without CHD and summarise maternal and environmental exposures, child characteristics and outcomes. Participants and non-participants will be compared using demographic and clinical characteristics collected during BH1 to assess potential selection bias. Continuous variables will be summarised using means and standard deviations or medians and interquartile ranges, as appropriate, and categorical variables using frequencies and percentages.

Outcomes derived from the standardised developmental assessments will be analysed among children with CHD. Mean performance will be compared with published normative expectations using appropriate one-sample tests, with effect sizes and 95% confidence intervals reported. Established thresholds will be used to describe developmental outcomes relative to normative expectations. Differences according to sex and CHD classification will also be examined.

Associations between preconceptional and antenatal maternal and environmental exposures and child outcomes will be examined using regression models appropriate to each outcome. Outcomes will include birth outcomes, physical development, parent-reported developmental outcomes, neurodevelopment (NEPSY-II), language (BPVS-3) and academic achievement (BAS3). Candidate exposures and potential confounders will be prespecified based on previous BH1 findings and existing literature. Multivariable models will be restricted according to the available sample size to reduce overfitting.

Parent-reported executive function (BRIEF-2) and behavioural and emotional outcomes (SDQ) will be compared between children with and without CHD. For outcomes available in both groups, analyses will examine whether associations between maternal and environmental exposures and child outcomes differ according to CHD status, where supported by the available sample size. Among children with CHD, associations between relevant sociodemographic, CHD-related and clinical factors and developmental outcomes will also be examined.

Analyses will use available data for each outcome, with sample sizes reported accordingly. Patterns and reasons for missing data will be examined and reported, and appropriate methods for handling missing data will be determined based on the extent and nature of missingness. The potential for multiple testing will be considered when interpreting findings and determining the final analytical approach. Findings will be interpreted considering effect estimates, 95% confidence intervals and consistency across related developmental domains. Analyses will be conducted using IBM SPSS Statistics 30.

## Discussion

This protocol describes the NI BabyHearts 2 Study (BH2), which builds on the original NI BabyHearts (BH1) case-control study through the longitudinal follow-up of children with and without CHD. By integrating prospectively collected maternal exposure data with developmental assessments, clinical information and administrative data linkage, BH2 provides a unique opportunity to investigate the contribution of early environmental factors to later child health and development.

### Strengths

This study has several notable strengths. The population-based design of BH1, which captured nearly all children diagnosed with CHD across Northern Ireland between 2014 and 2017, provides a strong foundation for longitudinal follow-up. High levels of consent for future recontact and continued engagement of families with paediatric cardiology services facilitate follow-up of children with CHD. The inclusion of children with and without CHD and integration of prospectively collected maternal exposure data, parental questionnaires, standardised developmental assessments and administrative data linkage provide a multidimensional approach to examining child outcomes. Active involvement of parents and key stakeholders throughout study design, implementation and dissemination further enhances the relevance and potential impact of the research.

### Limitations

Nevertheless, several limitations should be acknowledged. Attrition is expected given the time elapsed since BH1 and may be greater among children from the original control group, who do not have continued engagement with paediatric cardiology services. Differential participation may introduce selection bias, which will be assessed by comparing participants and non-participants using characteristics collected during BH1. Reliance on parental proxy reports may introduce reporting bias, although standardised developmental assessments and administrative and clinical data provide complementary sources of outcome information. The number of participants available for analyses of rare exposures and CHD subgroups may limit statistical power and the precision of effect estimates. Generalisability beyond the Northern Ireland population may also be limited; however, the population-based cohort and longitudinal design provide a valuable platform for future collaboration and pooled analyses with other cohorts.

### Conclusion

The NI BabyHearts 2 Study will provide new evidence on the relationship between preconceptional and antenatal environmental factors and later health and developmental outcomes. By examining children with and without CHD, the study will help determine whether associations between early environmental exposures and later outcomes are specific to children with CHD or reflect broader determinants of child development. Findings may inform clinical care and service provision, strengthen preconceptional and antenatal guidance, and contribute to public health strategies aimed at improving long-term outcomes for children with CHD and their families.

## Data Availability

No datasets were generated or analysed for the purposes of this Study Protocol. Data generated during the NI BabyHearts 2 study will not be made publicly available because of participant confidentiality and applicable ethical and data governance requirements. Access to study data may be considered upon reasonable request following study completion and will be subject to the necessary ethical, governance and data-access approvals.

https://doi.org/10.17605/OSF.IO/M9AS7

## Acknowledgements

The authors would like to thank the children and families participating in the NI BabyHearts studies for their continued involvement in the research. We thank the members of the PPIE Advisory Group and Children’s Heartbeat Trust for their contributions to the design and conduct of NI BabyHearts 2. In particular, we thank Joanne McCallister and Joan Aiken from Children’s Heartbeat Trust for their contributions as members of the PPIE Advisory Group and to the development of the successful funding application to Northern Ireland Chest Heart and Stroke. We also thank Dr Colette Duncan for her contribution to data collection through the administration of developmental assessments with participating children. The authors acknowledge the support of the Royal Belfast Hospital for Sick Children, the Belfast Health and Social Care Trust and the Honest Broker Service in facilitating the study and data linkage processes.

## Supporting Information

S1 Appendix. Full NI BabyHearts variable list by domain and source order. S2 Appendix. Developmental assessment measures and subtests.

